# SARS-CoV-2 Omicron variant escapes neutralization by vaccinated and convalescent sera and therapeutic monoclonal antibodies

**DOI:** 10.1101/2021.12.13.21267761

**Authors:** Nariko Ikemura, Atsushi Hoshino, Yusuke Higuchi, Shunta Taminishi, Tohru Inaba, Satoaki Matoba

**Affiliations:** Department of Cardiovascular Medicine, Graduate School of Medical Science, Kyoto Prefectural University of Medicine, Kyoto, Japan; Department of Infection Control and Laboratory Medicine, Graduate School of Medical Science, Kyoto Prefectural University of Medicine, Kyoto, Japan

## Abstract

The novel SARS-CoV-2 variant, Omicron (B.1.1.529) contains about 30 mutations in the spike protein and the numerous mutations raise the concern of escape from vaccine, convalescent sera and therapeutic drugs. Here we analyze the alteration of their neutralizing titer with Omicron pseudovirus. Sera of 3 months after double BNT162b2 vaccination exhibite ∼27-fold lower neutralization titers against Omicron than D614G mutation. Neutralization titer is also reduced in convalescent sera from Alpha and Delta patients. However, some Delta patients have relatively preserved neutralization activity up to the level of 3-month double BNT162b2 vaccination. Omicron escapes from the cocktail of imdevimab and casirivimab, whereas sotrovimab that targets the conserved region to prevent viral escape is effective to Omicron similarly to the original SARS-CoV-2. The ACE2 decoy is another modality that neutralize the virus independently of mutational escape and Omicron is also sensitive to the engineered ACE2.

---

The novel variant B.1.1.529 was detected in Botswana on November 11^th^ and spread rabidly and globally, then the World Health Organization (WHO) classified as Omicron variant of concern (VOC) on November 26^th^. Omicron has 26 to 32 mutations, 3 deletions and one insertion in the spike protein. Among them, 15 mutations are located in the receptor binding domain (RBD) (1). Spike mutations could enhance transmissibility and induce immune evasion (2). Compared to previous variants, Omicron contains numerous mutations in the spike and they could dramatically alter the characteristics of SARS-CoV-2. According to the routine surveillance data from South Africa, Omicron has higher transmission and risk of reinfection due to immune evasion (1). In addition, multiple mutations in the RBD have the impact on the escape from therapeutic monoclonal antibodies even in the cocktail treatment.

At first, we generated the pseudotyped virus harboring the spike of Omicron (https://covdb.stanford.edu/page/mutation-viewer/#omicron) and analyzed neutralizing activity of BNT162b2 vaccine and convalescent sera. Virus neutralization assay with vaccine sera obtained from 8 persons vaccinated with the Pfizer-BioNTech vaccine BNT162b2 showed ∼27-fold lower neutralization titers against Omicron than D614G mutation (parental virus). We also collected the convalescent sera before or after the pandemic of Delta variant. Convalescent sera before Delta pandemic (December 2020 through January 2021) showed ∼47-fold reduction and those of Delta pandemic (August 2021 through October 2021) exhibited ∼17-fold reduction. Importantly, among 11 patients during Delta pandemic, 5 patients’ sera exhibited less than 10-fold reduction and similar neutralization activity with those of 3 months after double BNT162b2 vaccination (Figure 1).

**Figure 1.**
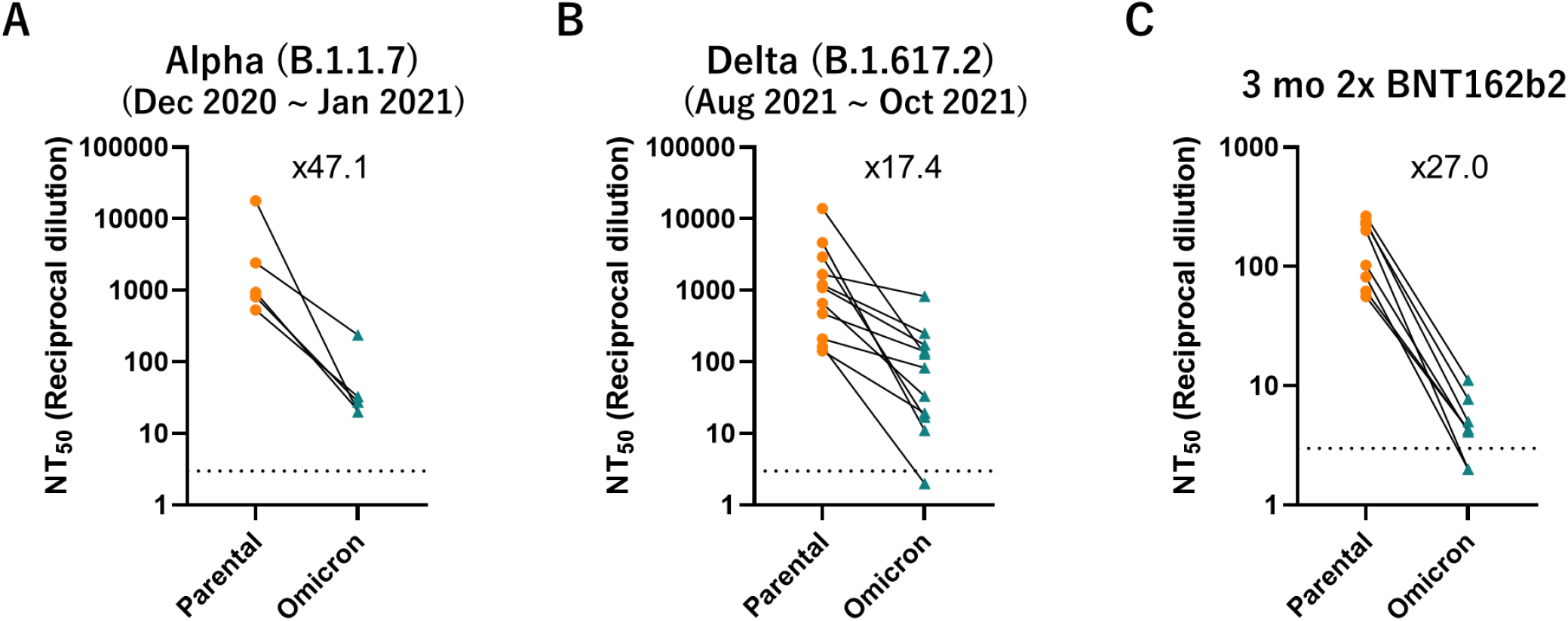
Neutralization assay of Omicron pseudovirus. The reciprocal dilution of 50% virus neutralization (NT50) was determined for sera of 3-month double BNT162b2 vaccination (A), convalescent sera from Alpha (B) and Delta (C). Dotted lines indicate detection limit.

We also evaluated the efficacy of neutralizing drugs developed for Wuhan strain. Currently the cocktail of imdevimab and casirivimab and sotrovimab were approved in Japan. The cocktail almost completely failed to neutralize Omicron, whereas sotrovimab showed the preserved neutralization (Figure 2A). We previously reported the therapeutic potential of the engineered ACE2 independently of mutational escape of virus (3). The virus variants escaping from ACE2 decoy should have the limited binding affinity to the native ACE2 receptors, which results in diminished or infectivity. As expected, the engineered ACE2 neutralized Omicron slightly better than Wuhan (Figure 2B).

**Figure 2.**
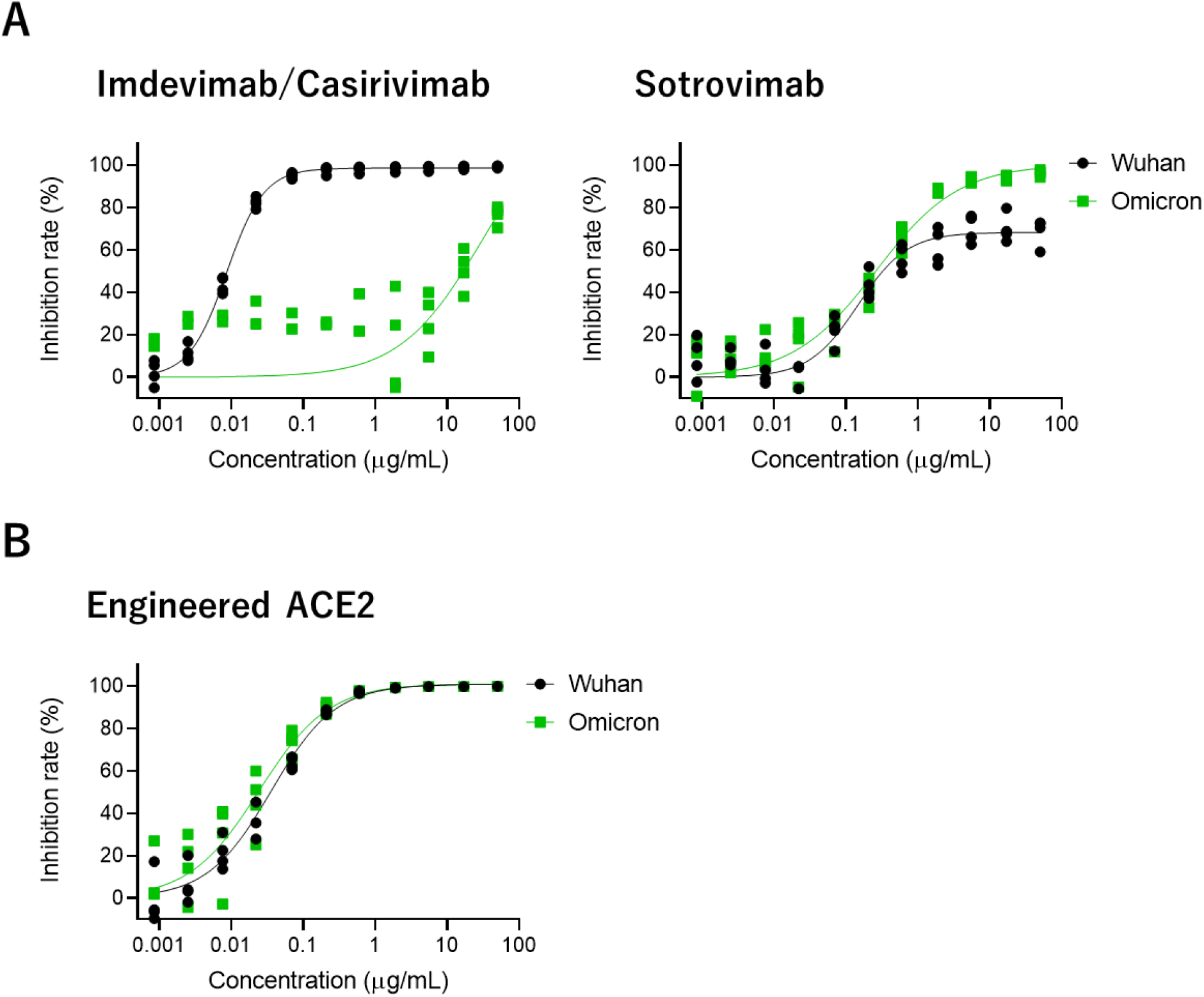
Neutralization assay for monoclonal antibodies and ACE2 decoy. Neutralization efficacy of the cocktail of imdevimab and casirivimab and sotrovimab against Omicron pseudovirus (A) and the engineered ACE2 (B). The indicated concentrations of casirivimab and imdevimab were applied in a 1:1 ratio.

Omicron contains 15 mutations in the RBD and escape from vaccine, convalescent sera, and some therapeutic antibodies. Although the partial cross neutralization against Omicron was observed in some patients during the Delta pandemic, it is required to develop the vaccine specific to Omicron to effectively prevent the Omicron pandemic. For therapeutic strategy to overcome the virus mutation, some antibodies are designed to target the conserved region to prevent the escape. However, Omicron obtain mutations even in that area (G339D, S371L, S373P, S375F), indicating the limitation to be free from the viral mutation. On the other hand, the engineered ACE2 successfully neutralized Omicron and could be the alternative approach to combat COVID-19 beyond the viral escape.

## Methods

### Ethics statement

Blood samples were obtained from hospitalized adults with PCR-confirmed SARS-CoV-2 infection and vaccinated individuals who were enrolled in a prospective cohort study approved by the Clinical Research Review Committee in Kyoto Prefectural University of medicine (ERB-C-1810-2, ERB-C-1949-1). All human subjects provided written informed consent.

### Cell culture

Lenti-X 293T cells (Clontech) and its derivative, 293T/ACE2 cells were cultured at 37 °C with 5% CO2 in Dulbecco’s modified Eagle’s medium (DMEM, WAKO) containing 10% fetal bovine serum (Gibco) and penicillin/streptomycin (100 U/ml, Invitrogen). All the cell lines were routinely tested negative for mycoplasma contamination.

### Protein synthesis and purification

Monoclonal antibodies and engineered ACE2 were expressed using the Expi293F cell expression system (Thermo Fisher Scientific) according to the manufacturer’s protocol. Fc-fused proteins were purified from conditioned media using the rProtein A Sepharose Fast Flow (Cytiva), respectively. Fractions containing target proteins were pooled and dialyzed against phosphate buffered saline (PBS).

### Pseudotyped virus neutralization assay

Pseudotyped reporter virus assays were conducted as previously described (3). A plasmid coding SARS-CoV-2 Spike was obtained from addgene #145032 and Omicron’s mutations and ΔC19 deletion (with 19 amino acids deleted from the C terminus) was cloned into pcDNA4TO (Invitrogen) (4). Spike-pseudovirus with a luciferase reporter gene was prepared by transfecting plasmids (OmicronΔC19, psPAX2-IN/HiBiT (5), and pLenti firefly) into LentiX-293T cells with Lipofectamine 3000 (Invitrogen). After 48 hr, supernatants were harvested, filtered with a 0.45 μm low protein-binding filter (SFCA), and frozen at –80 °C. The 293T/ACE2 cells were seeded at 10,000 cells per well in 96-well plate. HiBit value-matched Pseudovirus and three-fold dilution series of serum or therapeutic agents were incubated for 1 hr, then this mixture was administered to ACE2/293T cells. After 1 hr pre-incubation, medium was changed and cellular expression of luciferase reporter indicating viral infection was determined using ONE-Glo™ Luciferase Assay System (Promega) in 48 hr after infection. Luminescence was read on Infinite F200 pro system (Tecan). The assay of each serum was performed in triplicate, and the 50% neutralization titer was calculated using Prism 9 (GraphPad Software).

## Data Availability

All data produced in the present study are available upon reasonable request to the authors.

## Acknowledgements

This study has been performed with the support of Medical Research and Development (AMED), Research Program on Emerging and Re-emerging Infectious Diseases under 21fk0108465h0001 (A.H.). We would like to thank Junichi Takagi (Osaka University) for the synthesis of monoclonal antibodies and engineered ACE2.

